# Human and zebrafish data corroborate gene x gene interactions in ameliorating Duchenne muscular dystrophy disease progression

**DOI:** 10.64898/2026.11.01.26367015

**Authors:** Veronica J. Vieland, Geremy T. Lerma, Sang-Cheol Seok, Megan A. Waldrop, Leigh M. Gabel, Diane M. Dunn, Kevin M. Flanigan, Robert B. Weiss

**Affiliations:** Department of Pediatrics The Ohio State University, Columbus OH, USA; Department of Statistics (Emerita), The Ohio State University, Columbus OH, USA; Mathematical Medicine LLC, Chicago Il, USA; Department of Molecular Genetics, The Ohio State University, Columbus OH, USA; Center for Gene Therapy, The Abigail Wexner Research Institute at Nationwide Children’s Hospital, Columbus, Ohio, USA; Departments of Pediatrics The Ohio State University, Columbus, Ohio, USA; Departments of Neurology, The Ohio State University, Columbus, Ohio, USA; Department of Human Genetics, University of Utah, Salt Lake City, Utah, USA

**Author notes:** Correspondence to: VJ Vieland, Mathematical Medicine LLC 3800 N Lake Shore Dr Chicago IL (US).

**Keywords:** Duchenne muscular dystrophy, United Dystrophinopathy Project, gene x gene interaction, zebrafish, *MAN1A1*, *ACTN3*

## Abstract

Duchenne muscular dystrophy (DMD) is an X-linked recessive disorder involving progressive muscle tissue loss with replacement by fat and fibrotic tissue, leading in most cases to loss of ambulation (LOA) by early to mid-adolescence. We have previously reported the largest DMD genome-wide analysis to date, using N = 419 patients from the United Dystrophinopathy Project (UDP) whose primary *DMD* mutations are presumed to preclude production of residual dystrophin. We found strong evidence for 6 modifiers of LOA, 4 of which have been validated in zebrafish. Here we follow up by investigating possible gene-gene interactions both in the UDP and in zebrafish. We find strong evidence of interaction effects for 6 gene pairs in the UDP. We were able to test for interaction in zebrafish for one of these pairs, *MAN1A1* and *ACTN3*, finding strong evidence of an interaction effect with apparent rescue of muscle organization to nearly wild-type levels.

## Introduction

This report extends earlier findings [1, 2] by considering gene x gene (GxG) interaction effects among putative modifiers of Duchenne muscular dystrophy (DMD) in a cohort of 419 DMD cases from the multisite United Dystrophinopathy Project (UDP) [3-5]. DMD is an X-linked recessive disorder involving progressive muscle tissue loss with replacement by fat and fibrotic tissue. Patients typically become reliant on wheelchairs by early to mid-adolescence, but there is considerable variation in age at loss of ambulation (LOA), even among individuals whose primary *DMD* mutations are presumed to preclude any residual dystrophin production. Corticosteroid administration increases LOA by 2-3 years.

We previously reported strong evidence for association with LOA for the genes *ETAA1, NCALD* (or *KLF10*, see [2]), *GALNTL6, MAN1A1, ADAMTS19* and *PARD6G* [1]. Follow-up using a zebrafish DMD model appears to have confirmed a role for 4 out of 5 of these genes (*etaa1a*/*etaa1b, galntl6, man1a1, adamts19*) as modifiers [6]. (*PARD6G* was not a candidate for assay in the fish, and *adamts17* was tested as *ADAMTS19* is lost in zebrafish.) In a subsequent paper we considered gene x environment (GxE) interactions, focusing on interactions with corticosteroid administration [2]. We found compelling evidence of interactions with steroids in pathways regulating TGF-β bioavailability (*KLF10, LTBP4, THBS1, CD40*) and evidence against steroid interaction in the remaining 8 genes we considered, providing confirmatory evidence that genotypic variation within genes in the TGF-β pathway contributes to overall variability in DMD progression at least in part by modifying the efficacy of steroid administration, but also supporting the finding that the remaining modifiers on our list do not affect LOA by modulating steroid response. Here we present evidence of GxG interactions both in the UDP cohort and in the zebrafish model.

## Materials and Methods

The UDP cohort and molecular methods have been described in detail elsewhere [1] and are briefly recapped in Supplement§1. The phenotype is the Ordinary Time to Event (OTE) residual from a Weibull survival analysis [7, 8], which represents how unexpected is an individual’s LOA given the individual’s steroid status (Y/N). We continue to use the same statistical methodology tailored specifically to GWAS in the modest sample size available to us. Briefly, we originally developed a time-to-event version of the Posterior Probability of Linkage Disequilibrium (PPLD) [7-9] and used it for the initial GWAS. The PPLD is on the probability [0,..,1] scale, and we established heuristics for concluding moderate or strong evidence for trait-marker association (PPLD ≥ 0.1 and PPLD ≥ 0.4, respectively) using simulations. We then extended the statistical framework to cover GxE interactions, using the Posterior Probability of GxE Interaction (PPI_GxE_), in which we ask whether genotypic effects at a given SNP depend on steroid use [2].

Here we extend the underlying PPI_GxE_ framework to GxG interactions (PPI_GxG_), asking for each SNP pair whether the genotypic effect on LOA at one locus depends upon genotype at the other. See Supplement(§2) for additional details. Following the framework of our GxE work, we start with the 6 SNPs with strong evidence of association (PPLD ≥ 0.4) from our initial screen, plus a set of 6 candidate SNPs identified in other studies (see [1]), for a total of 66 pair-wise tests, and we report all results with moderate to strong evidence of GxG interaction (PPI_GxG_ ≥ 0.1).

Supplement(§3) provides the full list of SNPs and their associated genes. Simulations suggested that under the null hypothesis of “no interaction,” we would expect to see < 1 false positive signals using PPI_GxG_ ≥ 0.1 as our threshold; see Supplement(§4).

Estimates of mean OTE were obtained by maximizing the underlying log_10_ likelihood ratio (LR), with Support Intervals (SIs) displaying the lower and upper bounds on the estimate such that log_10_LR is within 2 log_10_ units of the maximum. SIs tell us the range of estimates that are approximately as well supported by the data as the maximum likelihood estimate itself, with the choice of 2 being conventional, and roughly equivalent under some regularity conditions to the choice of 95% for confidence intervals, although the interpretation is quite different.

The zebrafish experiments used a CRISPR-based screening approach, with birefringence intensity as the phenotype. Birefringence imaging is a technique that allows for the visualization of organized muscle tissue, such as seen in wild-type (WT), and the absence of birefringence seen in DMD mutant fish that is due to muscle disorganization [10]. The *dmd*^*ta222a*^ [11, 12] zebrafish strain was used to test pair-wise interactions. In brief, progeny of heterozygous *dmd*^*ta222a*^ adult zebrafish were microinjected with equal molar amounts of *modifier* sgRNA duplexes (IDT) and Cas9 protein (QB3-Berkeley, Cas9-NLS Purified Protein [2.5mg]), 0.1% Phenol Red, and 0.2M KCl, or raised as uninjected controls. All fish were raised to 4 days post-fertilization, processed for birefringence imaging [10], and then genotyped via HRMA [13] and PCR to identify if mutagenesis of the gene(s) of interest occurred via CRISPR/Cas9 and to validate *dmd* mutation status, respectively. See Supplement(§5) for additional details. Note that analysis of the *dmd-man1a1* crispant was previously reported in [6].

## Results

Table 1 shows results of the interaction analysis in the UDP data. There were 6 SNP-pairs with PPI_GxG_ ≥ 0.1, of which 5 had PPI_GxG_ ≥ 0.4.

**Table 1.** All PPI_GxG_ ≥ 0.1 in the UDP data (N = 419)

| **SNP #1** | **SNP #2** | **Gene 1** | **Gene 2** | **PPI<sub>GxG</sub>** |
|---|---|---|---|---|
| rs10077875 | rs1883832 | *ADAMTS19* | *CD40* | 1.00 |
| rs10077875 | rs28357094 | *ADAMTS19* | *SPP1* | 1.00 |
| rs1815739 | rs10499096 | *ACTN3* | *MAN1A1* | 1.00 |
| kgp9098261 | rs710160 | *TCTEX1D1* | *LTBP4* | 0.54 |
| rs1883832 | rs1815739 | *CD40* | *ACTN3* | 0.45 |
| rs10077875 | rs1358596 | *ADAMTS19* | *GALNTL6* | 0.27 |

SNPs were associated with genes based on the *in silico* pipeline described in detail in [1]. We note that of the 8 SNPs on this list, only 3 (*ADAMTS19*, *MAN1A1, GALNTL6*) showed strong evidence of association in our previous GWAS, with the remainder included based on results from other studies. This highlights the complex nature of these associations and the need for care in using non-replication to rule out a modifier role for any given gene.

To follow up in the zebrafish, we looked for the gene pair with the highest PPI_GxG_ such that both gene transcripts are well expressed in larval zebrafish, making them good candidates for knockdown experiments. This left us with the pair *ACTN3* and *MAN1A1* (both of which had showed strong evidence against interaction with steroid exposure [2], which is not further considered here). Since there are two orthologs of *ACTN3* in the fish, *actn3a* and *actn3b*, we considered *man1a1* interactions with each ortholog in turn.

We obtained PPI_GxG_ = 1.0 for interactions between *man1a1* and both *actn3* orthologs. Figure 1a shows effects on mean birefringence, making clear that there is a synergistic (greater than additive) effect of knocking down both genes at once. Mutating only one gene, *man1a1, actn3a* or *actn3b*, results in gains of 7.5, 3.2 and 5.3 in average birefringence, respectively. Under an additive model we would therefore expect increases of 7.5+3.2=10.7 (*actn3a, man1a1*) and 7.5+5.3=12.8 (*actn3b, man1a1*). But mutating either gene in the presence of the *man1a1* mutation adds 21.8 (*actn3a*) or 20.7 (*actn3b*) to the mean. For reference, mean refringence is 100.0 (SI 98.7-101.3) for WT, 45.5 (43.1-47.8) for *dmd*, and 67.3 (63.6-71.0), 66.2 (61.8-70.5) for the *dmd*-*man1a1-actn3a* and -*actn3b* double crispants, respectively. Figure 2 shows representative birefringence images for each of the conditions shown in Figure 1a. Visually, the double crispants appear to restore muscle organization to nearly WT levels. Of interest is the fact that in WT fish there is also a strong interactive effect, but in the opposite direction, with the double crispant fish showing more than additive *reductions* in birefringence (see Supplement(§7)).

**Figure 1.**
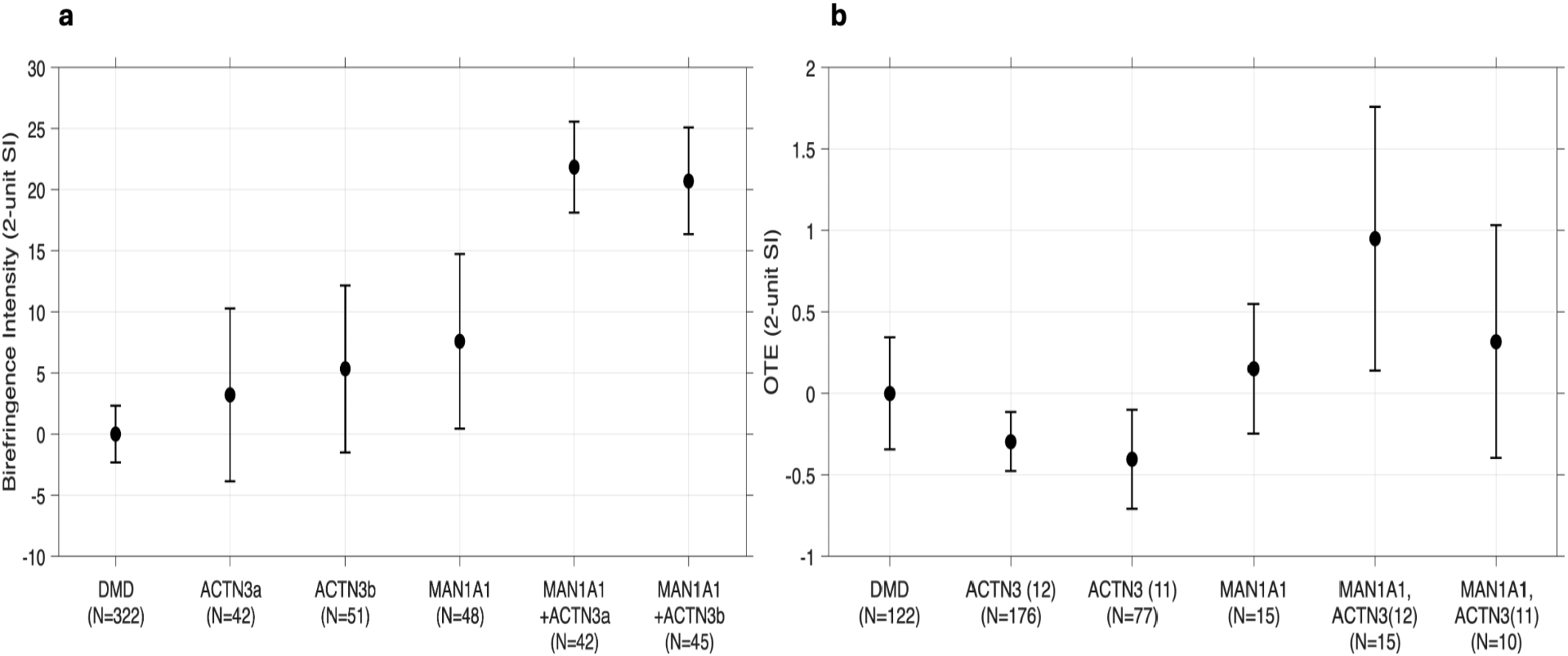
(a) Mean birefringence intensity in the zebrafish and (b) mean OTE in UDP data. Shown here is (a) mean normalized (to WT) birefringence intensity for non-crispant *dmd* fish, and for *dmd* fish mutated for each of the indicated genes as named along the x-axis, shown as differences from DMD means, along with the 2-unit Support Intervals; (b) mean OTE in the UDP data for each two-locus genotype, in standard deviation units and displayed as differences from the mean in DMD, along with 2-unit Support Intervals. Letting the common SNP allele be ‘2’ and the rare allele ‘1’ at each locus, the naming conventions are as follows, with the *ACTN3* genotype shown first: *DMD* = 22x22; *ACTN3*(12) = 12x22, *ACTN3*(11) = 11x22; *MAN1A1* = 22x12; *MAN1A1, ACTN3*(12) = 12x12; *MAN1A1, ACTN3*(11) = 11x12. Note that the y-axis units are different between the two subplots; note also that we lack sufficient data to analyze the *MAN1A1* 11 homozygote.

**Figure 2.**
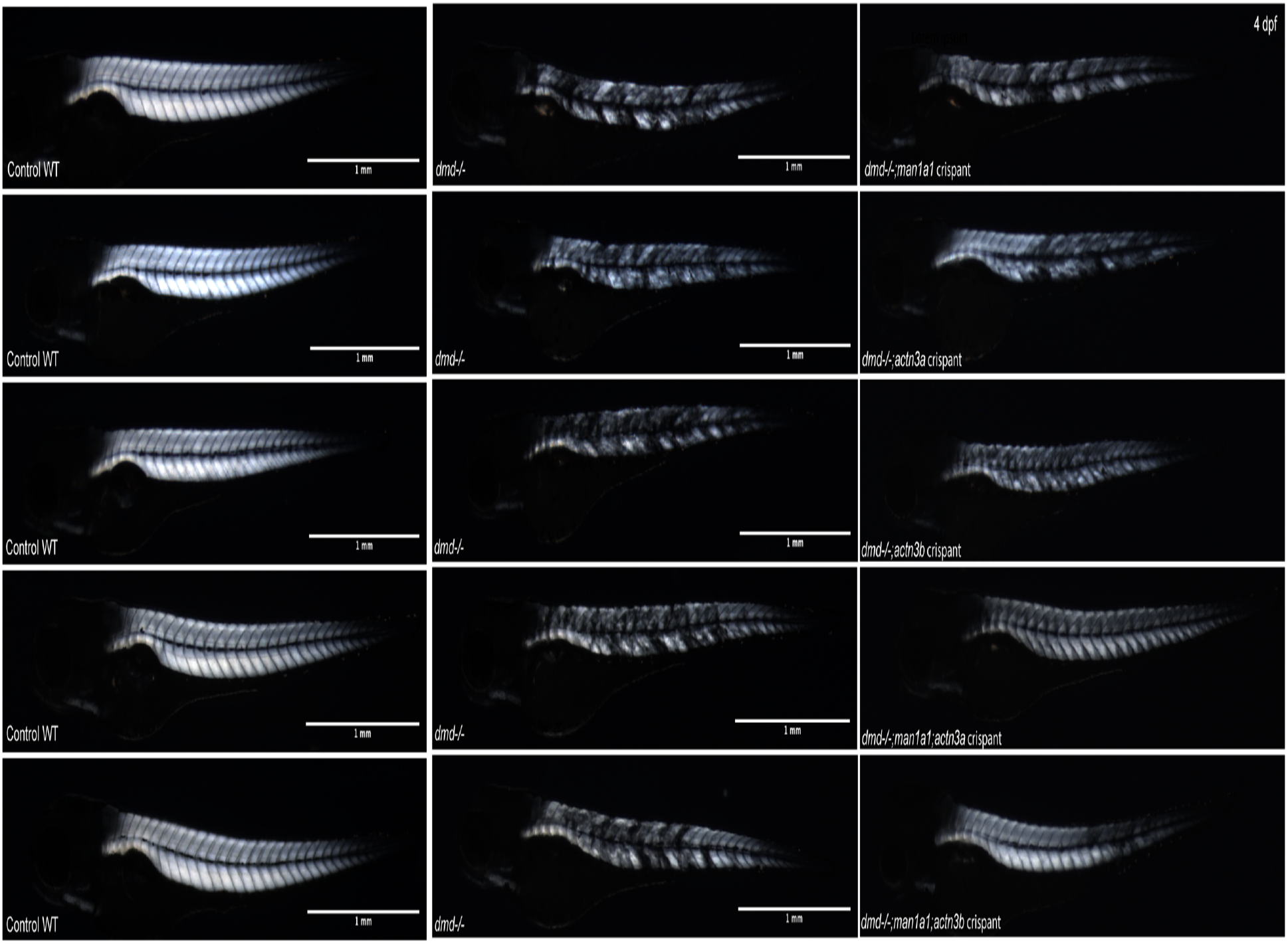
Representative birefringence zebrafish images for each of the conditions shown in Figure 1a.

In the original UDP GWAS [1] we obtained PPLD = 0.62 for *MAN1A1* but evidence against association for *ACTN3*: that is, the effect of *MAN1A1* on LOA was manifest even when we did not consider *ACTN3*, but the reverse was not true. For this gene pair we are now asking “Does the effect of *ACTN3* genotype depend on genotype at *MAN1A1*?” and the PPI_GxG_ = 1.0 is telling us that it almost certainly does. Note that there are only 2 individuals in the UDP homozygous for the rare *MAN1A1* allele, both of whom are homozygous for the common allele at *ACTN3;* these individuals are dropped from the analyses (see Supplement(§2)).

Figure 1b shows two-locus allelic effects on mean OTE in the UDP data in standard deviation units. There appears to be little if any effect of *ACTN3* genotype on OTE for the *MAN1A1* common homozygote, consistent with our original finding of evidence against association for this SNP. Referring to individuals with two-locus genotype 22, 22 as *DMD* (carrying mutations in *DMD* but not in either *ACTN3* or *MAN1A1*), we see that for *ACTN3* the means are 0.30 (*ACTN3*(12), where “(12)” indicates *ACTN3* genotype) and 0.41 (*ACTN3*(11)) lower than *DMD*, while the mean is 0.15 higher than *DMD* for *MAN1A1*. Under an additive model, we would therefore expect the mean increase in OTE due to variants in both genes to be −0.15 (*ACTN3*(12)), and −0.26 (*ACTN3*(11)). But in fact, the increases are 0.95 (*ACTN3*(12)), 0.32 (*ACTN3*(11)), which is higher than expected in both cases, by greater than one standard deviation for *ACTN3*(12). Unfortunately, in the absence of individuals carrying *ACTN3*(11) and *MAN1A1*(11), we cannot know how strong the effect would be in two-locus rare-allele homozygotes. (The PPI_GxG_ also allows for interaction effects on standard deviations; see Supplement(§6) for additional discussion for both the fish and UDP data.)

## Discussion

It is noteworthy that a pair of genes identified as showing strong evidence of interacting genotypic effects on LOA in a human data set would also show clear interactive effects on birefringence in a CRISPR knock-down in *dmd* zebrafish. These results highlight the utility of our zebrafish assays for rapid validation of human genetic findings. Our findings are all the more striking since we lack a mechanistic explanation of why these two genes would be interacting to influence the *dmd* phenotype; but clearly they do, showing large synergistic effects not only in *dmd* fish but also (in the opposite direction) in WT fish. Finally, the apparent rescue of muscle organization to nearly WT levels that we see in the *actn3/man1a1* mutated *dmd* zebrafish at 4 days post-fertilization is striking, and warrants further investigation.

## Data Availability

Data analyzed during the current study are available in the dbGaP repository at https://dbgap.ncbi.nlm.nih.gov/beta/study/phs003680.v1.p1/#study.

## Data Availability

The UDP data analyzed during the current study are available in the dbGaP repository at https://dbgap.ncbi.nlm.nih.gov/beta/study/phs003680.v1.p1/#study.

## ACKNOWLEDGEMENTS

The authors wish to acknowledge the many members of the United Dystrophinopathy Project consortium who participated in the enrollment of subjects in the original historical UDP database. We thank Sharon Amacher for her guidance and her support of the zebrafish experiments, and her entire laboratory for input and advice regarding the zebrafish work. We thank the Ohio State Zebrafish facilities staff for excellent zebrafish care, Danielle Pvirre for technical assistance, Dr. James Jontes for sharing equipment, Dr. Thomas Gallagher for qPCR advice, Dr. Jared Talbot for imaging advice, and Drs. Mayssa Mokalled and Dana Klatt Shaw for advice on CRISPR-based mutagenesis. We thank Carson Richardson for statistical discussions

## AUTHOR CONTRIBUTIONS

VJV, GTL, SCS, KMF and RBW conceived and designed the study. VJV, GTL, SCS, KMF, RBW, MAW, LMG and the UDP Investigators acquired and/or analyzed the data. VJV, GL, SCS drafted the manuscript, which was reviewed and revised by all of the named authors.

## COMPETING INTERESTS

The authors declare no competing interests. ETHICAL APPROVAL

The research involving human subjects has been performed in accordance with the Declaration of Helsinki and was approved by the Nationwide Children’s Hospital Institutional Review Board (IRB) under approval 0502HSE046. Informed consent was obtained from all subjects.

## References

1. Flanigan KM, Waldrop MA, Martin PT, Alles R, Dunn DM, Alfano LN, et al. A genome-wide association analysis of loss of ambulation in dystrophinopathy patients suggests multiple candidate modifiers of disease severity. Eur J Hum Genet. 2023;31(6):663–73.

2. Vieland VJ, Seok SC, Waldrop MA, Gabel LM, Dunn DM, Flanigan KM, et al. Gene x environment interaction analysis confirms genetic modifier effects on steroid efficacy via TGF-beta pathway in Duchenne muscular dystrophy. Eur J Hum Genet. 2026;34(7):964–71.

3. Flanigan KM, Dunn DM, von Niederhausern A, Soltanzadeh P, Gappmaier E, Howard MT, et al. Mutational spectrum of DMD mutations in dystrophinopathy patients: application of modern diagnostic techniques to a large cohort. Hum Mutat. 2009;30(12):1657–66.

4. Flanigan KM, Dunn DM, von Niederhausern A, Soltanzadeh P, Howard MT, Sampson JB, et al. Nonsense mutation-associated Becker muscular dystrophy: interplay between exon definition and splicing regulatory elements within the DMD gene. Hum Mutat. 2011;32(3):299–308.

5. Soltanzadeh P, Friez MJ, Dunn D, von Niederhausern A, Gurvich OL, Swoboda KJ, et al. Clinical and genetic characterization of manifesting carriers of DMD mutations. Neuromuscul Disord. 2010;20(8):499–504.

6. Lerma G, Ryhlick KR, Carraher OM, Beljan JC, Amacher SL. Validation of Duchenne muscular dystrophy candidate modifiers using a CRISPR-Cas9-based approach in zebrafish. bioRxiv. 2025.

7. Vieland VJ, Seok S-C, Stewart WCL. A new linear regression-like residual for survival analysis, with application to genome wide association studies of time-to-event data. PLoS One. 2020;15(5):e0232300.

8. Vieland VJ, Seok SC. The PPLD has advantages over conventional regression methods in application to moderately sized genome-wide association studies. PLoS One. 2021;16(9):e0257164.

9. Huang Y, Vieland VJ. Association statistics under the PPL framework. Genet Epidemiol. 2010;34(8):835–45.

10. Berger J, Sztal T, Currie PD. Quantification of birefringence readily measures the level of muscle damage in zebrafish. Biochem Biophys Res Commun. 2012;423(4):785–8.

11. Granato M, Eeden FJMv, Schach U, Trowe T, Brand M, Furutani-Seiki M, et al. Genes controlling and mediating locomotion behavior of the zebrafish embryo and larva. Development. 1996;123(1):399–413.

12. Li M, Hromowyk KJ, Amacher SL, Currie PD. Muscular dystrophy modeling in zebrafish. Methods Cell Biol. 2017;138:347–80.

13. Talbot JC, Amacher SL. A streamlined CRISPR pipeline to reliably generate zebrafish frameshifting alleles. Zebrafish. 2014;11(6):583–5.

