## Supplemental Material for "Human and zebrafish data corroborate gene x gene interactions in ameliorating Duchenne muscular dystrophy disease progression"

### 1. The UDP cohort

The UDP data used here have been previously described in [1, 2]. In brief, LOA was defined by full-time wheelchair use (requiring wheelchair use within the home) and recorded to the nearest month when available, and otherwise to the nearest half-year of age. Glucocorticoid treatment (Steroid Y) was defined as use of any steroid regimen for more than 6 months, beginning at least 6 months prior to loss of ambulation. Untreated (Steroid N) was defined as never exposed, exposed to any steroid regimen for less than 6 months, or with treatment onset less than 6 months prior to loss of ambulation. Genotyping methods and cleaning protocols are also described in [1]. The sample size is N = 419.

This sample was screened to include only subjects with true loss-of-function mutations within *DMD*, excluding genotypes known to be associated with milder disease severity (such as deletion of exons 3–7, deletion of exon 45, or nonsense mutations within the exon 23–42 region [3]). Based on available knowledge, we initially included individuals with duplication of exon 2 (Dup2) mutations; it was subsequently shown that some Dup2 carriers display milder phenotypes [3]. For this reason we repeated all analyses dropping the Dup2 individuals. There are 12 such individuals in the UDP dataset. When dropping DUP2 individuals (N = 407), results were similar, with one additional $PPI_{GxG}$ = 0.14 between *ACTN3* and *NCALD*.

### 2. Statistical Methods

Here we extend the underlying gene-by-environment interaction statistic, $PPI_{GxE}$, described in detail in [2] to consider gene-by-gene interactions using the $PPI_{GxG}$. $PPI_{G\times G}$ is a function of the likelihood ratio (LR) comparing two hypotheses, $H_1$ and $H_2$. While $PPI_{G\times E}$ was developed to answer to the question “Does the effect of steroid exposure on LOA depend on SNP genotype?”, $PPI_{G\times G}$ answer to the question “Does the effect of genotype at SNP1 on LOA depend on genotype at SNP2?”. Let the genotypes be represented by 11, 12 and 22 (where "1" is the minor allele) for each SNP. Let the genotypic mean and standard deviation (s.d.) of LOA be $\mu_{i,j}, \sigma_{i,j}$ for the $i^{th}$ genotype of SNP1 and the $j^{th}$ genotype of SNP2. Supp Table 1 shows the parameterization of the two-locus model.

**Supp Table 1** Parameterization of the GxG model

| | | **SNP2** | | | |
|---|---|---|---|---|---|
| **SNP1** | **genotype** | **11** | **12** | **22** | |
| | 11 | $\mu_{11,11}$ ($\sigma_{11,11}$) | $\mu_{11,12}$ ($\sigma_{11,12}$) | $\mu_{11,22}$ ($\sigma_{11,22}$) | $\mu_{11,*}$ ($\sigma_{11,*}$) |
| | 12 | $\mu_{12,11}$ ($\sigma_{12,11}$) | $\mu_{12,12}$ ($\sigma_{12,12}$) | $\mu_{12,22}$ ($\sigma_{12,22}$) | $\mu_{12,*}$ ($\sigma_{12,*}$) |
| | 22 | $\mu_{22,11}$ ($\sigma_{22,11}$) | $\mu_{22,12}$ ($\sigma_{22,12}$) | $\mu_{22,22}$ ($\sigma_{22,22}$) | $\mu_{22,*}$ ($\sigma_{22,*}$) |
| | | $\mu_{*,11}$ ($\sigma_{*,11}$) | $\mu_{*,12}$ ($\sigma_{*,12}$) | $\mu_{*,22}$ ($\sigma_{*,22}$) | $\mu_{*,*}$ ($\sigma_{*,*}$) |

Under $H_1$, the effects on LOA of genotype at SNP1 and genotype at SNP2 are assumed to be independent of one another, that is, the impact of SNP1 genotype is assumed to be the same regardless of SNP2 genotype. Under $H_2$, the impact of the SNP1 genotype is allowed to vary with SNP2 genotype. Supp Table 2 shows the parameterization of the two hypotheses.

**Supp Table 2** Parameterization of $H_1$, $H_2$

| | **11** | **12** | **22** |
|---|---|---|---|
| **$H_1$** | | | |
| **11** | $\mu_{11,11}, \sigma_{11,11}$ | $\mu_{11,12}, \sigma_{11,12}$ | $\mu_{11,22}, \sigma_{11,22}$ |
| **12** | $\mu_{11,11} + \alpha_{12}, \sigma_{11,11} \times \beta_{12}$ | $\mu_{11,12} + \alpha_{12}, \sigma_{11,12} \times \beta_{12}$ | $\mu_{11,11} + \alpha_{12}, \sigma_{11,22} \times \beta_{12}$ |
| **22** | $\mu_{11,11} + \alpha_{22}, \sigma_{11,11} \times \beta_{22}$ | $\mu_{11,12} + \alpha_{22}, \sigma_{11,12} \times \beta_{22}$ | $\mu_{11,11} + \alpha_{22}, \sigma_{11,22} \times \beta_{22}$ |
| **$H_2$** | | | |
| **11** | $\mu_{11,11}, \sigma_{11,11}$ | $\mu_{11,12}, \sigma_{11,12}$ | $\mu_{11,22}, \sigma_{11,22}$ |
| **12** | $\mu_{11,11} + \alpha_{12,11}, \sigma_{11,11} \times \beta_{12,11}$ | $\mu_{11,12} + \alpha_{12,12}, \sigma_{11,12} \times \beta_{12,12}$ | $\mu_{11,22} + \alpha_{12,22}, \sigma_{11,22} \times \beta_{12,22}$ |
| **22** | $\mu_{11,11} + \alpha_{22,11}, \sigma_{11,11} \times \beta_{22,11}$ | $\mu_{11,12} + \alpha_{22,12}, \sigma_{11,12} \times \beta_{22,12}$ | $\mu_{11,22} + \alpha_{22,22}, \sigma_{11,22} \times \beta_{22,22}$ |

These hypotheses can be succinctly represented as

$H_1$: $\alpha_{12,11} = \alpha_{12,12} = \alpha_{12,22},\ \beta_{12,11} = \beta_{12,12} = \beta_{12,22}, \alpha_{22,11} = \alpha_{22,12} = \alpha_{22,22},\ \beta_{22,11} = \beta_{22,12} = \beta_{22,22}$

$H_2$: $\alpha_{12,11} \neq \alpha_{12,12} \neq \alpha_{12,22},\ \beta_{12,11} \neq \beta_{12,12} \neq \beta_{12,22}, \alpha_{22,11} \neq \alpha_{22,12} \neq \alpha_{22,22},\ \beta_{22,11} \neq \beta_{22,12} \neq \beta_{22,22}$,

where "≠" means only that any of the equalities need not hold. This gives us the likelihood ratio

$$LR_{G\times G} = \frac{P(data \mid H_2)}{P(data \mid H_1)} = \frac{\prod_{i=1}^{3}\prod_{x_j\in i}\Phi(x_j; \mu_{11,i}+\alpha_{12,i}, \sigma_{11,i}\times\beta_{12,i})}{\prod_{i=1}^{3}\prod_{x_j\in i}\Phi(x_j; \mu_{11,i}+\alpha_{12}, \sigma_{11,i}\times\beta_{12})} \times \frac{\prod_{i=1}^{3}\prod_{x_j\in i}\Phi(x_j; \mu_{11,i}+\alpha_{22,i}, \sigma_{11,i}\times\beta_{22,i})}{\prod_{i=1}^{3}\prod_{x_j\in i}\Phi(x_j; \mu_{11,i}+\alpha_{22}, \sigma_{11,i}\times\beta_{22})},$$

where *i* ranges over genotypes; *j* ranges over individuals with a given genotype; $\mu_{11,i}, \sigma_{11,i}$ are the mean, standard deviation for the *i^th^* genotype of genotype 11 of SNP1 (which serves as the baseline), and Φ is the normal probability density function. Note that we assume normality of LOA at the genotypic level but not at the population level. See [2] Supplemental Material for additional computational details.

Continuing the development of the statistical framework developed for our GWAS analysis [4], we define the Bayes ratio (BR) as

$$BR_{G\times G} = log_{10} \int LR_{GxG}(\boldsymbol{\gamma})\, f(\boldsymbol{\gamma})\, d\boldsymbol{\gamma}$$

where the single integral stands in for multiple integration over the vector $\boldsymbol{\gamma} = (\alpha_{12,11}, \alpha_{12,12}, \alpha_{12,22}, \alpha_{22,11}, \alpha_{22,12}, \alpha_{22,22}, \beta_{12,11}, \beta_{12,12}, \beta_{12,22}, \beta_{22,11}, \beta_{22,12}, \beta_{22,22})$.

Finally, we convert the BR onto the posterior probability scale via Bayes' theorem:

$$PPI_{GxG} = \frac{\pi \times 10^{BR_{GxG}}}{\pi \times 10^{BR_{GxG}} + (1-\pi)}$$

where $\pi$ is set to 0.0004, the same point-prior used in calculating the PPLD. Since the true prior probability of an interaction is unknowable, there is no empirical basis for preferring any one value of $\pi$ over another; this particular choice for $\pi$ is one of convenience only, allowing us to maintain the same heuristics for evaluating evidence strength that we developed for the PPLD.

These same statistical methods were applied to the zebrafish data, treating each gene as if it were a 2-allele locus (crispant/non-crispant).

We note that following procedures established in [2], when the number of individuals in any cell in the two-locus data table is < 3, we drop the corresponding individuals from the LR calculation. In the current application this leads to dropping the 2 individuals homozygous for the rare *MAN1A1* allele.

All $PPI_{G\times G}$ calculations were done in MATLAB Version: 24.2.0.2773142 (R2024b).

**3. List of SNPs used in this study**

**Supp Table 3.** The list of 12 SNPs used in this study.

| Associated Gene | SNP | MAF | PPLD | $PPI_{GxE}$ |
|---|---|---|---|---|
| ETAA1 | rs34263553 | 0.130 | 0.87 | 0.0005 |
| NCALD | rs72681143 | 0.106 | 0.77 | 0.0000 |
| GALNTL6 | rs1358596 | 0.186 | 0.42 | 0.0000 |
| MAN1A1 | rs10499096 | 0.053 | 0.62 | 0.0000 |
| ADAMTS19 | rs10077875 | 0.481 | 0.44 | 0.0000 |
| PARD6G | rs2061566 | 0.091 | 0.40 | 0.0001 |
| SPP1 | rs28357094 | 0.224 | 0.00 | 0.0000 |
| LTBP4 | rs710160 | 0.248 | 0.00 | 1.00 |
| THBS1 | rs2725797 | 0.142 | 0.00 | 1.00 |
| CD40 | rs1883832 | 0.266 | 0.00 | 0.0000 |
| ACTN3 | rs1815739 | 0.438 | 0.00 | 0.0000 |
| TCTEX1D1 | kgp9098261 | 0.141 | 0.00 | 0.0002 |

6 of these SNPs comprise all PPLDs ≥ 0.4 from our initial genome scan and the remaining 6 were identified in other studies (see [1] for details). PPLD and $PPI_{GxE}$ results are repeated from [1], [2], respectively, for reference.

**4. Null simulations**

In theory, if the effect of genotype at SNP #1 depends on genotype at SNP #2, then the reverse is also true. However, power to detect dependence can vary appreciably depending, upon other things, on discrepant minor allele frequencies between the two SNPs. For this reason, we analyze each SNP-pair twice, conditioning on each SNP in turn, and report the larger of the resulting $PPI_{GxE}$ statistics. Because this procedure will have some inflationary effects, we used simulations to assess the null distribution of the resulting statistic for the initial UDP analyses.

**Supp Table 4** shows the generating models used for these simulations, following procedures previously described in detail [2, 5]. In brief, for each replicate, 400 individuals were simulated with 2 randomly drawn minor allele frequencies from the minor allele frequency distribution in the DMD data set. An individual was simulated based on a random draw of $t_e(x_i)$ from an age-at-event (AE) distribution, as given by the generating parameters and incorporating left-truncation of the distribution to avoid negative ages, and an independent random draw of $t_o(x_i)$ from an age-at-observation (AO) distribution. AO was simulated under a negative binomial distribution with $r = 11, p = 0.4$. If $t_e(x_i) < t_o(x_i)$, the individual was considered uncensored with failure time $t_{fail}(x_i) = t_e(x_i)$; otherwise, the individual was considered censored with censoring time $t_{cens}(x_i) = t_o(x_i)$. We simulated 10,000 replicates, each analyzed twice (conditioning SNP2 on SNP1 and vice versa). There were 130/10,000 replicates (0.65%) with $PPI_{G\times G} \geq 0.1$. Thus out of our 66 pair-wise tests, we would expect to see < 1 false positive signals using PPI ≥ 0.1 as our threshold.

**Supp Table 4** Null simulation generating model

| | | SNP2 | | |
|---|---|---|---|---|
| | **genotype** | **11** | **12** | **22** |
| **SNP1** | 11 | 16.0 (2.0) | 15.0 (2.0) | 14.0 (2.0) |
| | 12 | 16.0 (2.0) | 15.0 (2.0) | 14.0 (2.0) |
| | 22 | 13.0 (2.0) | 12.0 (2.0) | 11.0 (2.0) |

Shown here is the model for the Steroid N group, which accounts for 30% of each replicate; parameters for the steroid Y group are obtained by adding 3 years to the mean and multiplying the standard deviation by 1.5, to reflect steroid effects in the real data.

### 5. Zebrafish methods

*Zebrafish Stocks and Husbandry* Adult zebrafish strains (Danio Rerio) were kept at 28.5°C on a 14h light/dark cycle and embryos were obtained by natural spawning or in vitro fertilization and staged as previously described [6]. The *dmd*$^{ta222a}$ strain [7, 8] has been maintained on the AB6 wild-type (WT) background for many generations.

*DNA extraction and dmd*$^{ta222a}$ *genotyping* Individual embryos and adult fin tissue were lysed in 50µl 1X ThermoPol Buffer (NEB) at 95°C for 10min, digested at 55°C for 1hr in 1mg/ml ProteinaseK (Thermofisher), followed by Proteinase K inactivation at 95°C for 10min. 1µl of DNA extract was used in 1 20µl PCR reaction with Taq polymerase. *dmd*$^{ta222a}$ genotyping PCR was carried out with forward (5'- CATACCCAAGGTTTCAAAGCA -3') and reverse (5'-TGCACTCGAGTGAAGCCACGTTTTT-3') primers . PCR reactions were then digested with DraI (20 units) at 37°C for 30 minutes and analyzed on a 2% agarose gel stained with Gel Red (Biotium) to distinguish *dmd* WT (140 bp product) and *dmd*$^{ta222a}$ (110 bp product) alleles.

*CRISPR/Cas9 mutagenesis* CRISPR/Cas9 design and mutagenesis were performed as previously described [9] [10]. crRNA guide RNA sequences were selected using either CHOPCHOP or IDT CRISPR Design Tool. Lyophilized Alt-R tracrRNA and crRNA gRNAs (IDT, Cat# 1072534) were prepared as previously described [9]. In short, tracrRNA (100µM) and crRNA (100µM) were mixed (50µM), annealed by heating to 95°C and gradual cooling to 25°C, diluted to 25µM and stored at -20°C until use. Cas9 protein (QB3-Berkeley, Cas9-NLS Purified Protein [2.5mg]) was diluted to 25uM and stored at -80°C. For injection, sgRNA duplexes and Cas9 protein were mixed 1:1 in equal molar amounts with 1M KCl (0.2M final concentration) and Phenol Red (0.1% final concentration). Progeny of *dmd*$^{ta222a}$ heterozygous adult incrosses were injected with 1nL of CRISPR/Cas9 solution at the one-cell stage and raised for phenotypic assays. All injected fish were genotyped via HRMA to identify whether the target region was mutagenized. All CRISPR guides are listed in Supplemental Table 5.

*High-resolution melt analysis* High-resolution melt analysis (HRMA) was conducted as previously described [11] to identify if mutagenesis had occurred at the target site. In short, primers were designed (Primer3) flanking the CRISPR target site. DNA was extracted from adult fin tissue or whole larvae as described above and 1ul was used as template in a 10µl HRMA reaction (BioRad 172-5112) in a CFX Duet Real-Time PCR System (#12016265) and analyzed using BioRad Precision Melt Analysis Software. All HRMA primers are listed in Supplemental Table 5.

**Supp Table 5** CRISPR guides and HRMA genotyping primers used in this study.

| Gene | CRISPR gRNA | HRMA Forward Primer (5'-3') | HRMA Reverse Primer (5'-3') |
|---|---|---|---|
| *man1a1* | CGTGTCCAGTGCGTCCACAA | CCCCACAGGATGCCTTCTCAAA | CACCCATTCGGTAGCGGCTTCA |
| *actn3a* | CTAAGGAGGGCTTGCTGCTG | GCTAGTGAAAATATTCAACTGACCT | AAAGAAGATTTCACCTGATGTGG |
| *actn3b* | AGGCTGCCCAAACCTGACAA | TGCTCAGTGAGTGTGTGTTGAT | TGCTACAGATGAAATCCAGAGC |

*Birefringence Imaging and Analysis* Birefringence was used to quantitatively assess dystrophy severity [12] as previously described [10]. Zebrafish were raised to 4 days post-fertilization (dpf), fixed overnight in 4% PFA, and rinsed with PBST 5 times for 10 minutes. A 35mm glass bottom dish was previously prepared for imaging by using a 1% agar solution and plastic mold [13] to create 'inserts' to hold fish for ease of imaging. Fish were individually placed into inserts for imaging. Using a Leica MZFLIII stereomicroscope equipped with polarizing filters, birefringence images were taken with a Zeiss AxioCam Hrc camera using AxioVision SE64 Rel 4.8 software. ImageJ was used to quantify mean gray values using the polygon selection tool to outline the entire trunk of the fish. Mean grey values are presented as a percentage of the average mean gray value of the WT siblings in their respective control group.

*Normalization* In order to remove potential confounding effects, within each clutch birefringence intensities were normalized with respect to the mean birefringence of WT fish assayed in the same experiment. This yields intensity as a percent of WT, as reported in Results.

We note that the double crispant for *actn3a* and *actn3b* leads to a significant reduction in birefringence intensity in WT (Supp Figure 1) which prevents testing in *dmd-/-* zebrafish.

**Supp Figure 1 Birefringence imaging of double *actn3a-actn3b* crispant fish**

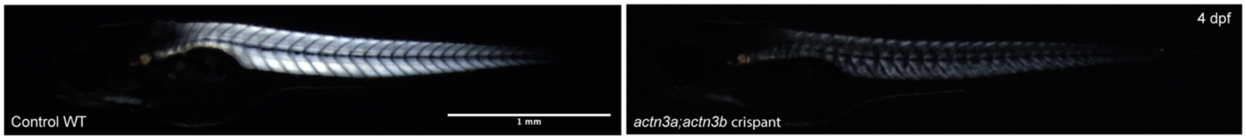


### 6. GxG effects on standard deviations in zebrafish and UDP

Our statistical model, $PPI_{G \times G}$, measures not only the changes in means but also in standard deviations. While "no interaction" corresponds to an additive model for the means, there is not a simple relationship to additivity for effects on standard deviations. Nevertheless, it is possible to estimate and plot the standard deviations and their SIs as we did for the means, as shown in Supp Figure 2.

**Supp Figure 2** Effects on standard deviations

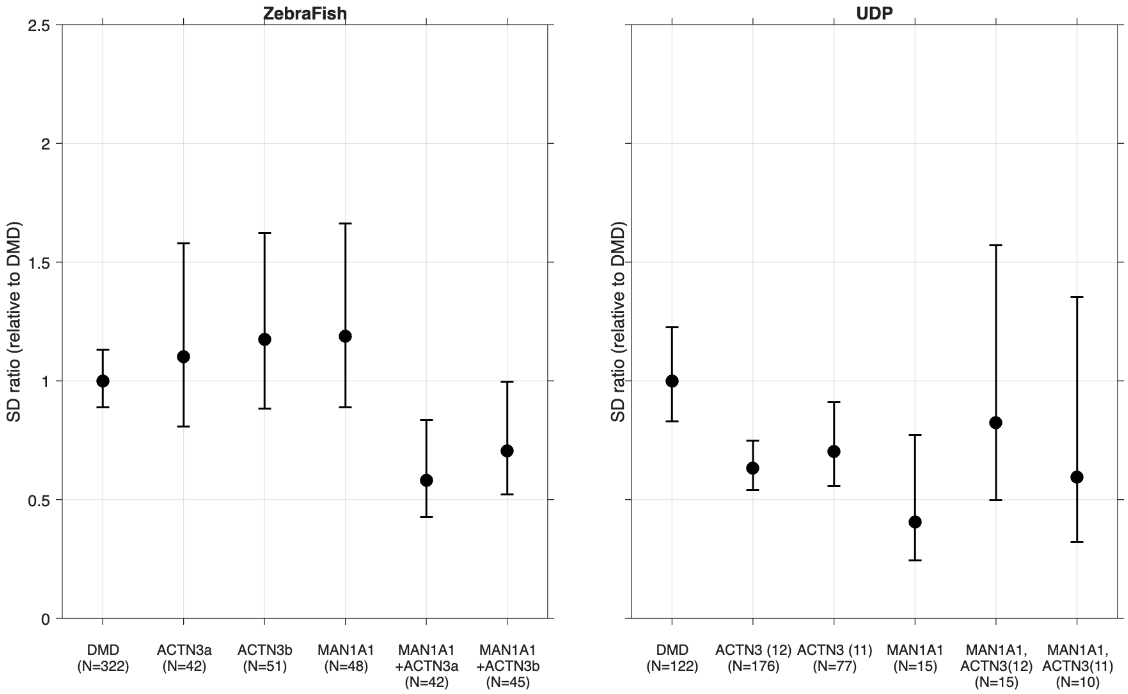


Shown here are standard deviations of (a) normalized birefringence intensity for each group of *dmd* fish and (b) standard deviations of OTE in the UDP DMD data set, along with 2 ($log_{10}$) unit Support Intervals. In each case results are plotted as ratios relative to DMD.

### 7. Knockdown effects in WT fish

While our focus in this paper is on interactive gene effects in Duchenne patients and in *dmd* fish, as mentioned in the main text, we also see an interactive effect in wildtype fish, but the effect goes in the oppositive direction as shown in Supp Figures 3 and 4. $PPI_{GxG}$ = 1.0 for both *actn3a* and *actn3b*, and in both cases, the double mutants show substantially *lower* birefringence intensity compared to the others.

**Supp Figure 3** Knockdown effects in fish on a WT background

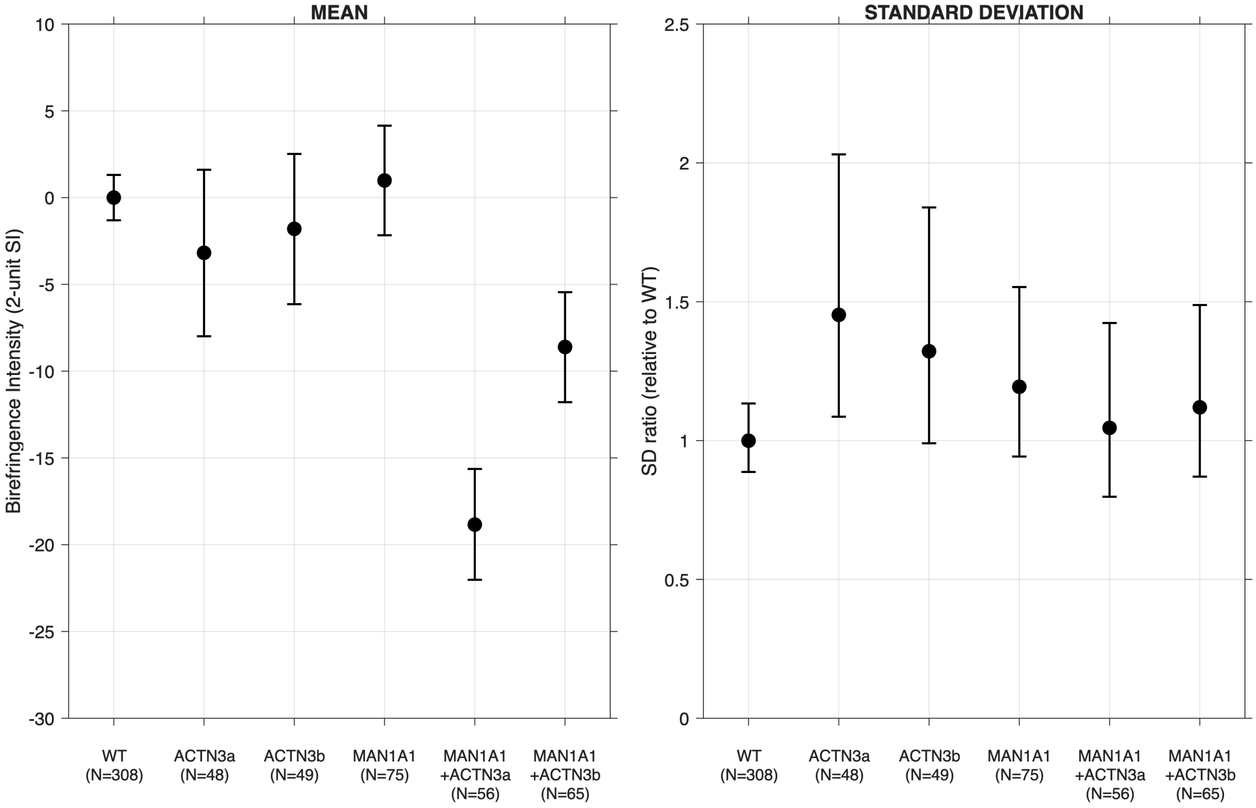

**Supp Figure 4 Birefringence intensity in WT fish**

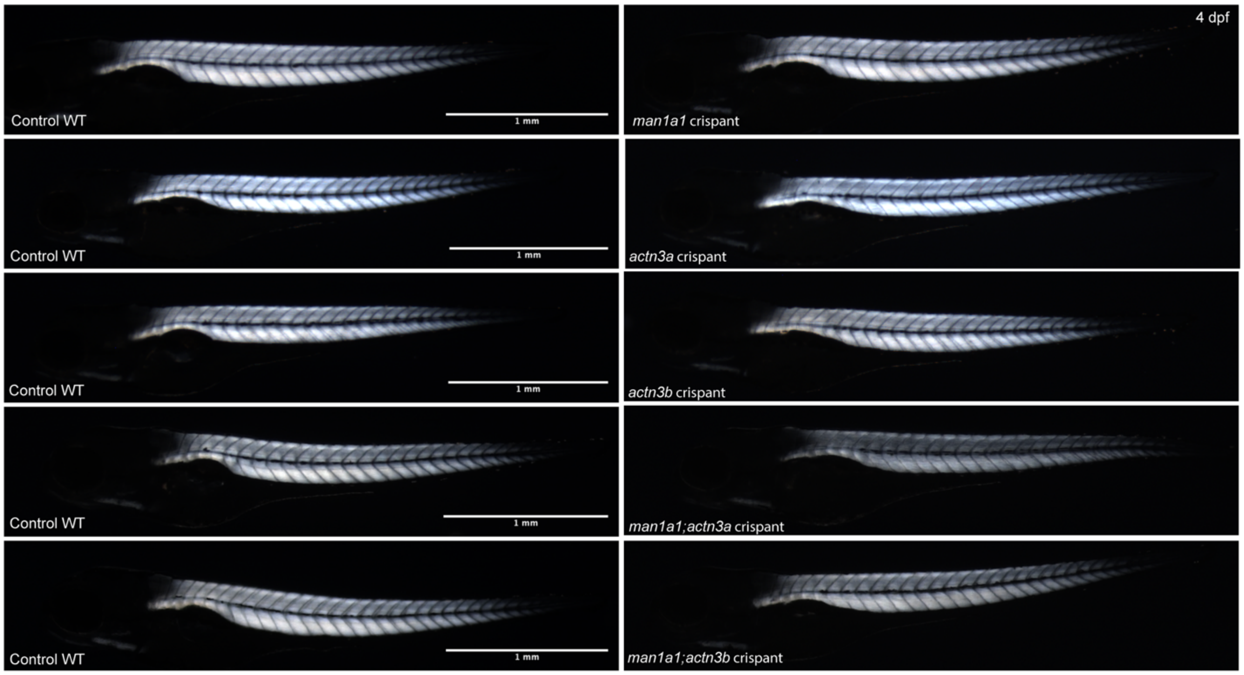


### References


1. Flanigan KM, Waldrop MA, Martin PT, Alles R, Dunn DM, Alfano LN, et al. A genome-wide association analysis of loss of ambulation in dystrophinopathy patients suggests multiple candidate modifiers of disease severity. Eur J Hum Genet. 2023;31(6):663-73.
2. Vieland VJ, Seok SC, Waldrop MA, Gabel LM, Dunn DM, Flanigan KM, et al. Gene x environment interaction analysis confirms genetic modifier effects on steroid efficacy via TGF-beta pathway in Duchenne muscular dystrophy. Eur J Hum Genet. 2026;34(7):964-71.
3. Zambon AA, Waldrop MA, Alles R, Weiss RB, Conroy S, Moore-Clingenpeel M, et al. Phenotypic Spectrum of Dystrophinopathy Due to Duchenne Muscular Dystrophy Exon 2 Duplications. Neurology. 2022;98(7):e730-e8.
4. Vieland V. J., Huang Y., Seok S-C., Burian J., Catalyurek U., O'Connell J., et al. KELVIN: a software package for rigorous measurement of statistical evidence in human genetics. Human Heredity. 2011;72(4):276-88.
5. Vieland VJ, Seok SC. The PPLD has advantages over conventional regression methods in application to moderately sized genome-wide association studies. PLoS One. 2021;16(9):e0257164.
6. Kimmel CB, Ballard WW, Kimmel SR, Ullmann B, Schilling TF. Stages of embryonic development of the zebrafish. Dev Dyn. 1995;203(3):253-310.
7. Granato M, Eeden FJMv, Schach U, Trowe T, Brand M, Furutani-Seiki M, et al. Genes controlling and mediating locomotion behavior of the zebrafish embryo and larva. Development. 1996;123(1):399-413.

8. Li M, Hromowyk KJ, Amacher SL, Currie PD. Muscular dystrophy modeling in zebrafish. Methods Cell Biol. 2017;138:347-80.
9. Klatt Shaw D, Mokalled MH. Efficient CRISPR/Cas9 mutagenesis for neurobehavioral screening in adult zebrafish. G3 (Bethesda). 2021;11(8).
10. Lerma G, Ryhlick KR, Carraher OM, Beljan JC, Amacher SL. Validation of Duchenne muscular dystrophy candidate modifiers using a CRISPR-Cas9-based approach in zebrafish. bioRxiv. 2025.
11. Talbot JC, Amacher SL. A streamlined CRISPR pipeline to reliably generate zebrafish frameshifting alleles. Zebrafish. 2014;11(6):583-5.
12. Berger J, Sztal T, Currie PD. Quantification of birefringence readily measures the level of muscle damage in zebrafish. Biochem Biophys Res Commun. 2012;423(4):785-8.
13. Megason SG. In toto imaging of embryogenesis with confocal time-lapse microscopy. Methods Mol Biol. 2009;546:317-32.
